# Analytical Validation of a Pan-Cancer NGS Assay for In-House Liquid Biopsy Testing: An International Multicenter Study

**DOI:** 10.1101/2024.10.17.24313324

**Authors:** Gaëlle Lescuyer, Alexandre Harlé, Hari Shankar Kumar, Pantelis Constantoulakis, Nicole Pfarr, Ellen Heitzer, Clémence Michon, Gianluca Russo, Ernst-Jan M Speel, Marie Piecyk, Marie Husson, Georgia Christopoulou, Eva-Maria Mayr, Mai-Lan Koppermann, Christophe Passot, Ricarda Graf, Anes Hadjadj Aoul, Violaine Bourdon, Hendrikus J Dubbink, Ronald van Marion, Imke Demers, Anne-Marie C Dingemans, Giancarlo Troncone, Francesco Pepe, Laura Muinelo-Romay, Ángel Díaz-Lagares, Aitor Rodriguez-Casanova, Ramón Manuel Lago Lestón, Deepak Pathak, Parth Shah, Romain V Parillaud, Oskar Martínez de Ilarduya, Jonas Behr, Alexis Rapin, Thomas Vetterli, Sanga Mitra Boppudi, Umberto Malapelle, Lea Payen-Gay

## Abstract

**Background:** Liquid biopsy (LBx) assays are transforming precision oncology by the screening of genomic alterations in cfDNA. These assays provide a less invasive alternative to tissue biopsies, which are not always feasible. Molecular pathology laboratories require LBx assays that detect variants at low allele frequencies using standardized methods.

**Methods:** This study evaluated the Hedera Profiling 2 ctDNA test panel (HP2) (Hedera Dx, Epalinges, Switzerland), a hybrid capture-based NGS assay for the detection of somatic alterations from cfDNA. Covering 32 genes, HP2 enables the detection of SNVs, Indels, Fusions, CNVs, and MSI status from a single DNA-only workflow. The analytical performance was assessed using reference standards and a diverse cohort of 137 clinical samples pre-characterized by orthogonal methods.

**Results:** In reference standards at 0.5% VAF, detection sensitivity and specificity for SNVs/Indels were 96.92% and 99.67%, respectively, and 100% each for Fusions. For MSI with VAFs of ≥1% and CNVs with VAFs of ≥ 2% both achieved 100% sensitivity.

**Conclusion:** This international, multicenter analytical performance evaluation study across a large number of hospital laboratories demonstrated high concordance of HP2 assay with orthogonal methods, confirming its significant potential as a highly sensitive, and efficient Pan-Cancer test for future decentralized LBx testing.

## Introduction

The advancements in next-generation sequencing (NGS) technologies have remarkably transformed cancer testing for an increasing number of tumors, enabling the potential to identify actionable somatic genomic alterations at very low variant allele frequencies with increased analytical sensitivity and specificity (1,2). The growing recognition of cancer-derived somatic mutation detection in advanced solid tumors resonates with the emphasis from European Society for Medical Oncology (ESMO) guidelines (3), American Society of Clinical Oncology (ASCO) provisional clinical opinion (4) and the recommendations provided by the National Comprehensive Cancer Network (NCCN) (5). These guidelines for clinical routine cancer management underscore the significance of plasma-based cell-free and circulating tumor nucleic acids testing, specifically within the domain of evidence-based consensus on circulating tumor DNA (ctDNA) analysis. In the era of cancer genomic profiling, liquid biopsy (LBx) (non-invasive approach) assays are increasingly being used in many scenarios where collecting a traditional tissue sample is painful, invasive or limited. Their applications span across advanced stage molecular profiling, early detection and screening, therapy selection, and monitoring for minimal residual disease and cancer recurrence. Despite the rapidly increasing integration of NGS systems in clinical practice, a notable void persists in the standardization and comprehensive validation of LBx assays tailored for pan-cancer applications. This limitation underscores a critical gap in the field, necessitating concerted efforts to establish rigorous protocols and validation frameworks to ensure the technical reliability and consistency of these innovative diagnostic testing across a spectrum of cancer types.

The analytical validation of an pan-cancer NGS LBx test is an essential step to ensure its reliability and accuracy (6). Moreover, validated procedures encompass evaluating the NGS LBx assay across diverse sample types and conditions to ascertain its resilience and consistency in varying scenarios. Furthermore, scrutinizing potential error sources and integrating stringent quality control measures are pivotal facets of the analytical validation process, safeguarding the overall precision and trustworthiness of the test outcomes (7). Beyond the meticulous analytical validation with commercial reference samples, the evaluation of analytical performance utilizing clinical samples of the pan-cancer NGS LBx test assumes a pivotal role in transitioning theoretical efficacy to tangible biological performance. Assessing sensitivity, specificity, and limit of detection with clinical samples establishes the test’s reliability in-vivo. Comparative analysis with orthogonal methods enhances its credibility, while rigorous validation of the bioinformatics pipeline ensures accurate and reproducible variant detection, crucial for identifying actionable mutations and guiding personalized oncology treatments.

In this context, Hedera Dx has developed a blood-based solution that streamlines the process of suggesting adequate therapy options for major cancers, with the goal of enabling hospital laboratories to run clinically actionable LBx assays in-house. The Hedera Profiling 2 ctDNA test panel (HP2) is a 32-gene assay (90 kb design size) optimized for detecting various genomic somatic alterations, including Single Nucleotide Variants (SNVs), Insertions and Deletions (Indels), Copy Number Variations (CNVs), splice variants, Gene Fusions and Microsatellite Instability (MSI). Employing a streamlined DNA-only single workflow, enhanced with the implementation of Unique Molecular Identifiers (UMIs) for molecular barcode error detection, this assay ensures sensitive identification of genomic alterations removing PCR duplicates and sequencing artifacts. The HP2 panel covers clinically relevant biomarkers and most promising molecular alterations included in major guidelines for most common solid tumors. The HP2 assay is intended for in vitro examination of blood plasma circulating free DNA (cfDNA) using targeted hybrid-capture NGS workflow, and specifically focuses on ESCAT (ESMO Scale of Clinical Actionability for molecular Targets) Evidence tier I - II genomic alterations. This product is currently labeled for Performance Studies Only (PSO), for the evaluation of actionable tumor biomarkers supporting clinical decision making strategy in non-small cell lung cancer (NSCLC), colorectal cancer (CRC), thyroid cancer, breast cancer (BC), melanoma, cholangiocarcinoma (CHOL), and gastrointestinal stromal tumor (GIST) (8,9) and other solid tumors.

The EMERALD Trial (NCT03778931) highlighted the importance of detecting *ESR1* activating mutations in ER+/HER2-advanced breast cancer, demonstrating that Elacestrant, a novel oral ER degrader, significantly benefited ESR1+ BC patients who progressed after endocrine therapy including CDK4/6 inhibitors (10,11). This led to EMA approval of Elacestrant in September 2023. *ESR1* mutations, detectable in up to 40% of ER+/HER2-advanced or metastatic BC, contribute to resistance against conventional endocrine therapy (12).

To address this need, the performance of HP2 for detecting *ESR1* mutations in LBx was successfully evaluated through a Quality in Pathology (QuIP) proficiency test. Simultaneously, the assay’s capability to detect *MET* exon 14 skipping mutations (METex14) in LBx for NSCLC was also assessed with QuIP. METex14, resulting from various genetic alterations, leads to overactive *MET* signaling pathway and tumor growth (13,14). The complexity of these mutations, with hundreds of distinct alterations reported, emphasizes the need for robust detection methods. This comprehensive evaluation by QuIP targets two critical genetic alterations with significant implications for cancer diagnostics and treatment strategies in BC and NSCLC.

In the present study, we outline a comprehensive analytical validation study for liquid-biopsy based HP2 assay. This involves utilizing commercially available, pre-characterized cfDNA or formalin-fixed, paraffin-embedded (FFPE) reference samples alongside with clinical plasma samples obtained either from a commercial biorepository or research samples through accredited labs (with patient consent) representing different solid tumors.

## Materials and methods

### Samples and nucleic acid isolation

#### Commercial reference samples

A total of 10 cfDNA, 2 FFPE and 1 gDNA commercially available reference standards were sourced from 4 different vendors (Supplementary Table S1).

#### Clinical plasma samples

Clinical plasma samples were obtained from 11 referral institutions involved in molecular testing. A total of 137 clinical specimens were used, from donors diagnosed with either NSCLC (N=98), Breast (N=12), CRC (N=9), Prostate (N=5) or other cancer types (N=8) (Supplementary Table S2). Samples were orthogonally tested for SNVs, Indels, Fusions, CNVs and MSI status using alternative NGS assays (based on targeted hybrid capture or amplicon techniques), either in cfDNA or in matched FFPE samples (Supplementary Table S3). Whole blood was either collected in Streck Cell-Free DNA BCT tubes (Streck, La Vista, NE, USA) or Cell-free DNA Collection tube (Roche Diagnostics GmbH, Mannheim, Germany) or EDTA tubes and plasma was collected by double centrifugation within 4-48 hours (for Streck and Roche tubes) and 6 hours for EDTA tubes according to the manufacturer’s guidelines. Plasma samples were stored at −80 °C until cfDNA extraction.

#### Clinical FFPE/gDNA samples

A total of 25 clinical FFPE/gDNA samples, from donors diagnosed with either NSCLC (N=5), CRC (N=8), Endometrium (N=5) or other cancer types (N=7) (Supplementary Table S4). These samples were used for the evaluation of MSI analysis performance only, in the context of limited clinical cfDNA specimens, and their MSI status was orthogonally determined through capillary electrophoresis methods.

#### cfDNA isolation from plasma

cfDNA was isolated from a minimum of 4 mL of plasma using commercial kits according to the manufacturer’s guidelines (see Supplementary materials and methods for details). Extracted cfDNA was quantitated using the Qubit dsDNA High-Sensitivity Assay Kit (Thermo Fisher Scientific, Waltham, MA, USA). Isolated cfDNA was stored at −20 °C.

#### DNA isolation from FFPE samples

DNA was isolated using QIAamp DNA FFPE Tissue kit (Qiagen, Hilden, Germany) according to the manufacturer’s guidelines. Extracted DNA was quantitated using the Qubit dsDNA Broad Range Assay Kit (Thermo Fisher Scientific, Waltham, MA, USA). Isolated DNA was stored at −20 °C.

### Sequencing library preparation and sequencing

Sequencing libraries were prepared with the Hedera Profiling 2 ctDNA test panel (HP2) assay (Hedera Dx, Epalinges, Switzerland) following the manufacturer’s instructions. Briefly, DNA was extracted from plasma or FFPE samples and 30 ng were used as input material. Adapters including a Unique Molecular Identifier (UMI) sequence were ligated and ligated DNA fragments were then purified using magnetic beads. Fragments were amplified by PCR with primers containing sample-specific Unique Dual Index (UDI) sequences; libraries were multiplexed in pools of 6 at equimolar concentration. Target regions were then enriched using hybrid-capture biotinylated probes overnight and purification with streptavidin-coated beads. The resulting enriched libraries were diluted and prepared for sequencing according to Ilumina’s guidelines (Illumina, San Diego, CA). Libraries were sequenced on Illumina NextSeq series (500/550/550Dx/1000/2000) or NovaSeq 6000 instruments using Paired-End (PE) 2x150 bp chemistry and 10 bp dual-index reads.

For FFPE and gDNA samples, a preliminary step of enzymatic fragmentation was applied with conditions that lead to fragment profiles similar to cfDNA. Briefly, in a single reaction including end-repair and A-tailing, DNA was fragmented using Hedera enzymatic fragmentation kit (Hedera Dx) following the manufacturer’s instructions. Subsequent steps mirrored those outlined above for cfDNA, including UMI adaptor ligation.

### Bioinformatics pipeline and data analysis

Sequencing data were analyzed using Hedera Prime^TM^ (version 1.9.1), a registered CE marked in vitro medical device software, for secondary and tertiary analysis. The Hedera Prime medical device includes an alignment module, a duplex consensus and SNVs/Indels calling module, a CNV detection module and additional PSO computational modules for Fusions detection and MSI classification. The Hedera Prime MSI module determines MSI status by analyzing 36 genomic simple repeat loci, classifying samples as Microsatellite Stable (MSS) or Microsatellite Instable (MSI-high) based on significant alterations in at least 6 loci from expected repeat lengths. Subsequent data analyses were performed with Python 3.10 (see Supplementary materials and methods for details).

### Analytical validation

The HP2 assay performance was evaluated using 160 individual DNA libraries derived from commercially available reference standards and clinical cfDNA samples, and sequenced on Illumina NextSeq and NovaSeq instrument series (see Supplementary materials and methods for details). Briefly, this analytical validation assessed the performance of SNVs, Indels and Fusions detection at 0.5% VAF, the performance of CNV detection at 0.5% and 2% VAF as well as the performance of MSI status determination on a range of *in-silico* dilutions mimicking 0% to 5% allele frequencies.

The reproducibility of SNVs, Indels and Fusions detection at 0.5% VAF was assessed using independent replicates of a same DNA library sequenced in two distinct runs and estimating the concordance of results within and between runs. The assay reproducibility was also assessed across independent laboratories and sequencing instruments, both in terms of concordance and observed VAF variations. Briefly, the reproducibility of SNVs and Indels detection across independent diagnostic laboratories (inter-laboratory concordance) was assessed using a same cfDNA reference standard including 8 SNVs and Indels at 1% spike-in frequency (SensID 5-Gene-Multiplex reference). Inter-instrument reproducibility was assessed for SNVs, Indels and Fusions detection using a same set of DNA libraries derived from reference standards at 0.5% spike-in frequency and sequenced on the NextSeq 550, NextSeq 2000 and NovaSeq 6000 instruments.

The impact of VAF on SNVs, Indels and Fusions detection was assessed using reference standards with spike-in frequencies at 0.1%, 0.25%, 0.5%, 1%, 2.5% and 5%. The impact of input DNA material on SNVs and Indels detection was assessed using reference standards at 0.5% spike-in frequency and a variation of the sequencing library preparation protocol using 10 ng, 20 ng, 30 ng and 50 ng of input DNA instead of the standard 30 ng.

### Round robin test / QuIP participation

The performance of HP2 for the detection of *ESR1* and METex14 mutations in LBx was assessed within the scope of a QuIP, 2024 proficiency test conducted at the Institute of Pathology - Technical University Munich (TUM, Munich, Germany). This rigorous assessment involved 39 participants for *ESR1* testing and 16 participants for METex14 testing, representing institutions from Germany, Austria, and Switzerland. Both certification assessment schemes encompassed 10 human plasma samples, each spiked-in with artificial cfDNA harboring specific variants manufactured by SensID GmbH (Rostock, Germany). For the *ESR1* scheme, the samples incorporated single or double mutations for 8 clinically relevant SNVs within exons 5 and 8 of *ESR1* gene. For the METex14 scheme, focused on single mutations falling within Intron 13, 14 or Exon 14 of the *MET* gene. Libraries were prepared using 30 ng input DNA with HP2 assay and sequencing results were analyzed with the Hedera Prime software.

### Clinical concordance

The concordance between the HP2 assay and orthogonal NGS assays was assessed in terms of SNVs, Indels, Fusions and CNV detection, as well as MSI status determination, using 137 clinical cfDNA specimens (Supplementary Table S3). SNVs/Indels reported with a VAF below 0.1% by orthogonal methods, as well as variants reported solely by HP2, were excluded.

## Results

### HP2 Assay - Panel overview and sequencing metrics

The HP2 assay was designed to detect SNVs and Indels in 32 genes, CNVs in 14 genes, and Fusion events in 9 genes (Supplementary Table S5). The panel also integrates METex14 and *EGFR* variant VIII as well as MSI detection through 36 MSI markers. Out of the 32 genes covered by the panel, it includes 18 of the 21 ESCAT level I genes recommended by ESMO for ctDNA analysis, representing approximately 86% coverage (3). The remaining are either frequently mutated genes or predictive biomarkers for solid tumors. The HP2 assay includes reagents for NGS library preparation and a dedicated web application, Hedera Prime for the sequencing data analysis.

From reference standards, a total of 160 libraries were generated (sequenced in 21 runs) which are used in the assay evaluation, generating in average 54.74±13.19M reads per library (8.40±2.14 Gb). Following UMI-based duplex consensus calling, this represented 1.19±0.27M aligned duplex consensus reads per library, with a mean coverage on target of 869.83±208.59x and a mean coverage on exons relevant for SNVs and Indels classified as ESCAT Level I for NSCLC of 1030.28±244.07x (Supplementary Table S6).

To assess the relevance of results obtained on reference standards for clinical settings, the coverage profiles obtained from 23 DNA libraries derived from Twist cfDNA reference standard were compared to those obtained from 14 clinical cfDNA samples. Each library was prepared using the HP2 kit with 30 ng input DNA and sequenced either on a NextSeq or NovaSeq 6000 instrument (Supplementary Table S7). Regions falling within exons including SNVs and Indels classified as ESCAT Level I for NSCLC showed higher coverage in both coverage on individual target exons was consistent between reference standards and clinical samples (R^2^=0.58, p<0.0001) (Fig. 1a, Fig. S1b). Low coverage was consistently observed on *FGFR3* exon 2, *EGFR* exon 1 and a portion of *ALK* exon 16 (maximum observed coverage below 260x in reference standards at 0.5% VAF), and three variants included in the Twist cfDNA reference standard falling on these regions were systematically excluded from all performance and concordance analyses.

**Fig. 1:**
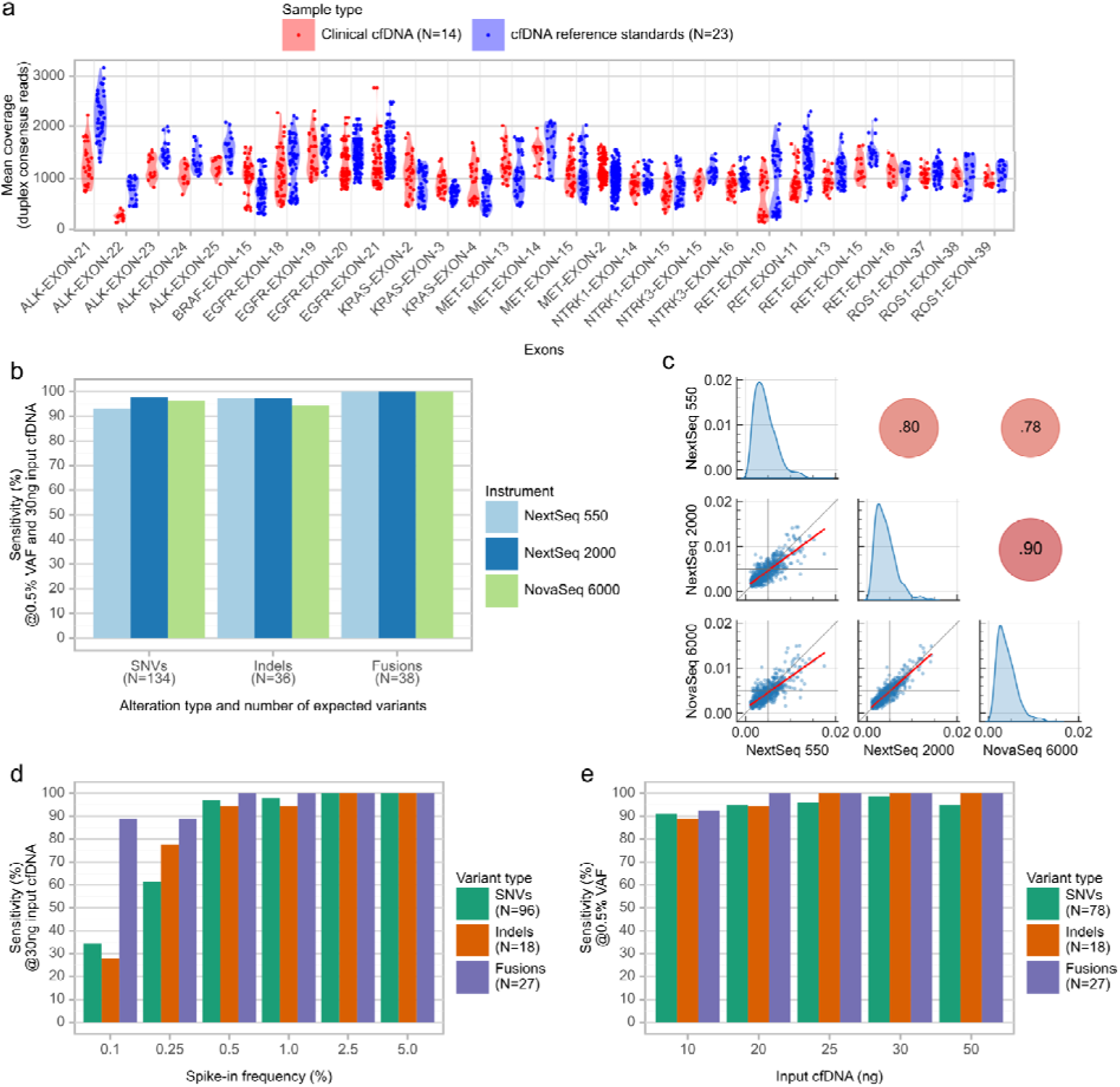
SNV, Indels and Fusions detection performance on cfDNA reference standards. **a.** Mean duplex consensus reads coverage per gene exon covered by the HP2 panel. Data points represent regions of 150bp. Data shown for exons relevant for ESCAT Level I mutations. **b.** Sensitivity obtained with a same set of DNA libraries derived from reference standards at 0.5% spike-in VAF and sequenced on different Illumina sequencers. Data shown for SNVs, Indels and Fusions relevant for ESCAT Level I mutations. **c.** Observed variant allele fractions (VAF) pair-plot for SNVs and Indels detected in a same set of DNA libraries derived from reference standards at 0.5% spike-in VAF and sequenced on different Illumina sequencers. Bottom right triangle: VAF shown for individual variants. Red line denotes a linear regression. Vertical and horizontal gray lines represent the 0.5% VAF. Diagonal: Kernel density estimates of VAF. Top left triangle: Pearson’s correlation coefficients. **d.** Analytical sensitivity obtained on reference standards with different spike-in frequencies using 30 ng of input DNA. **e.** Analytical sensitivity obtained on reference standards at 0.5% spike-in frequency using different input DNA quantities.

### Variant detection performance - SNVs, Indels and Fusions

#### Reference samples

Performance was assessed on reference standards at 0.5% spike-in frequency using 30 ng of input DNA (Table 1, Supplementary Tables S9-11), independently for SNVs, Indels and Fusions and for each following variant subset:

- ‘**All variants**’ represents the total number of tested variants falling within the HP2 panel target regions, which includes 2150 variants (1348 SNVs and 802 Indels) plus 54 Fusions.
- ‘**ESCAT Level I - All tumor types**’ represents the subset of ‘All variants’ which are termed as Level 1 (A/B/C) for any tumor type, and include 422 variants (336 SNVs and 86 Indels) plus 54 Fusions.
- ‘**ESCAT Level I - NSCLC**’ represents the subset of ‘All variants’ which are termed as Level 1 (A/B/C) for NSCLC, and include 287 variants (221 SNVs and 66 Indels) plus 54 Fusions.

**Table 1:** Analytical performance summary. Summary of the HP2 assay SNVs, Indels, Fusions CNVs detection, as well as MSI status determination sensitivity and specificity. SNVs and Indels results presented by variant subsets representing ESCAT classification. All results obtained using 30 ng input DNA. *: For SNVs and Indels, the reported VAF and supporting reads depth are based on duplex consensus reads, while for Fusions, these metrics are based on raw sequencing reads. **: MSI status determination performance reported at limit of detection based on *in-silico* dilutions.

| Alteration type | Spike-in frequency | Variant subset | Number positives / Number expected | Analytical sensitivity | Analytical specificity | Observed VAF / fold-change (median and range)* | Observed VAF / fold-change CV (mean±sd)* | Supporting reads depth (median and range)* |
| --- | --- | --- | --- | --- | --- | --- | --- | --- |
| SNVs | 0.5% | ESCAT Level I NSCLC | 220/221 | 99.55% | 99.70% | 0.38% (0.11-1.25%) | 35.4±13.2% | 8 (2-23) |
|  |  | ESCAT Level I (all tumor types) | 329/336 | 97.92% | 99.70% | 0.38% (0.11-1.95%) | 35.8±13.3% | 8 (2-28) |
|  |  | All variants | 1233/1348 | 91.47% | 99.70% | 0.38% (0.11-1.95%) | 31.7±18.7% | 7 (2-28) |
| Indels |  | ESCAT Level I NSCLC | 60/66 | 90.91% | 99.97% | 0.36% (0.13-0.80%) | 45.2±14.4% | 8 (2-22) |
|  |  | ESCAT Level I (all tumor types) | 80/86 | 93.02% | 99.97% | 0.36% (0.13-0.80%) | 42.3±14.3% | 8 (2-22) |
|  |  | All variants | 714/802 | 89.03% | 99.97% | 0.36% (0.10-1.41%) | 41.0±16.4% | 6 (2-25) |
| Fusions |  | All variants | 54/54 | 100.00% | 100.00% | 0.27% (0.06-1.98%) | 205.7±63.7% | 44 (18-170) |
| CNVs | 2.5% | N/A | 2/2 | 100.00% | 100.00% | 1.71x (1.65-1.77x) | N/A | N/A |
|  | 0.5% | N/A | 15/24 | 62.50% | 98.28% | 1.17x (1.14-1.57x) | 8.99±5.86% | N/A |
| MSI | 1.5%** | N/A | 1/1 | 100.00% | N/A | N/A | N/A | N/A |

The observed SNVs/Indels VAF on reference standards at 0.5% spike-in frequency ranged from 0.10% to 1.95% (0.37% median), with a mean CV of 40.25±13.77%, in line with manufacturer’s specifications (Supplementary Fig. S2).

#### Repeatability and reproducibility

The HP2 inter- and intra-assay reproducibility was assessed on two identical sequencing libraries prepared and sequenced independently, and including replicate DNA libraries from a same set of 4 reference standards (Supplementary Table S12). For variants classified as ESCAT Level I for NSCLC, the inter-assay concordance (i.e. percentage of variants consistently detected across the 2 libraries) reached 100.00% for SNVs, 92.59% for Indels and 98.26% for Fusions. For the same variants, the average intra-assay concordance (i.e. percentage of variants consistently detected across replicates within a same sequencing library) reached 100.00±0.00% for SNVs, 89.42±15.77% for Indels and 96.68±9.34% for Fusions (Supplementary Table S13).

The inter-laboratory reproducibility of the HP2 assay was assessed by analyzing a synthetic reference standard with a 1.0% spike-in VAF across six independent diagnostic laboratories (Table 2). Utilizing 30 ng of input DNA, the study revealed an average pairwise concordance of 87.5% and an average sensitivity of 93.75%. The minor discrepancy in variant detection was attributed to insufficient sequencing depth in one reference standard, further highlighting the assay’s exceptional robustness and precision across different laboratory settings.

**Table 2:**
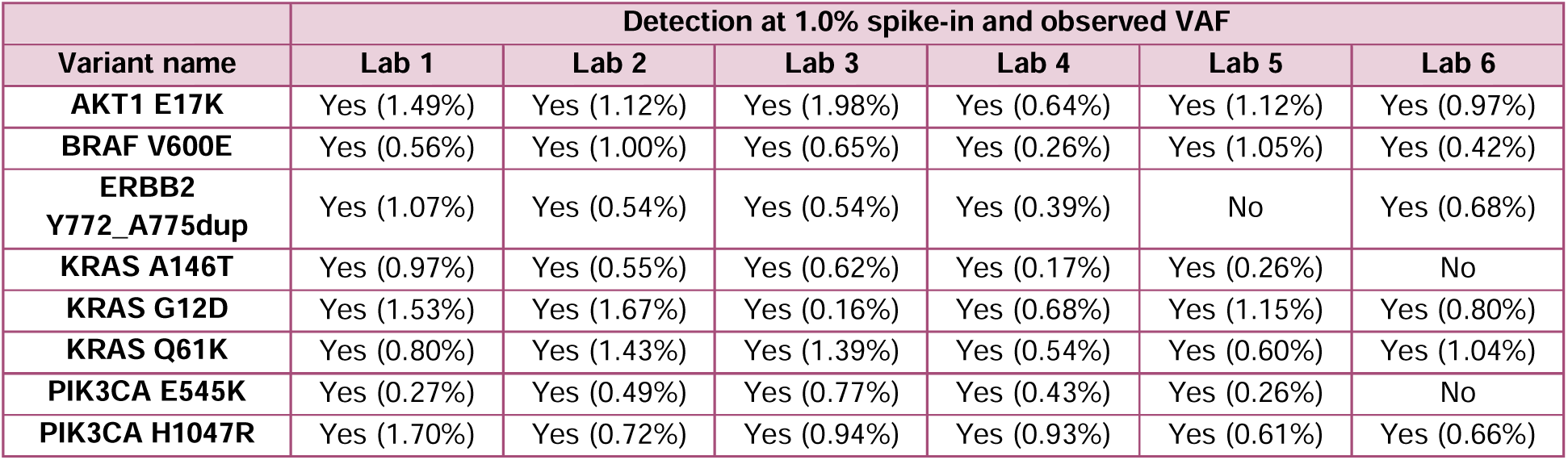
Inter-lab reproducibility on reference standards. Observed VAF at 1% spike-in frequency in reference standards processed in 6 independent laboratories. Results obtained using 30 ng input DNA.

The HP2 assay inter-instrument concordance was assessed on Illumina NextSeq 550, NextSeq 2000 and NovaSeq 6000 instruments using reference standards at 0.5% spike-in VAF using 30 ng of input DNA. Sequencing yield was highest on the NovaSeq 6000 instrument, with 0.24M more aligned duplex consensus reads obtained than on the NextSeq 550 in average (Supplementary Table S14). The assay showed a consistently high sensitivity for SNVs, Indels and Fusions across instruments (Fig. 1b), and a concordance of 93.86-96.52% on variants classified as ESCAT Level I for NSCLC (Supplementary Tables S15-16). The observed VAF was highly correlated between instruments (Fig. 1c).

#### Impact of VAF on variant detection

The impact of VAF on variant detection was assessed using the same set of reference standards at different spike-in frequencies (Supplementary Table S17). Although variants in libraries at low spike-in frequencies (i.e. below 0.5%) were detected with a lower sensitivity, the HP2 assay could detect variants in libraries with a spike-in frequency as low as 0.1% (Fig. 1d, Supplementary Fig. S3, Supplementary Table S18).

#### Impact of input DNA quantity on variant detection

The impact of input cfDNA quantity on variant detection was assessed using the same set of reference standards at 0.5% spike-in VAF (Fig. 1e, Supplementary Fig. S4, Supplementary Table S19-20). Libraries prepared from lower amounts of cfDNA (10 ng) showed fewer numbers of aligned duplex consensus reads while a larger number was obtained from 30 ng cfDNA and a large variability was found in libraries prepared from 50 ng cfDNA (Supplementary Fig. S5).

#### Round robin test / QuIP participation

The HP2 assay demonstrated excellent performance in the QuIP proficiency tests for both *ESR1* and METex14 mutation detection in LBx samples, presenting 100% accuracy. For *ESR1* mutations, HP2 successfully passed the test along with 85% of participants (33/39), accurately identifying 11 clinically relevant mutations across 10 samples (Supplementary Table S21). In the METex14 proficiency test, HP2 was among the 62.5% of successful participants (10/16), correctly detecting 8 clinically relevant *MET* mutations and two wild-type samples in a set of 10 blinded specimens. Notably, participants who did not pass either test were using other commercially available NGS liquid biopsy-specific assays, highlighting the superior performance of HP2 in these challenging detection scenarios.

#### Clinical samples - SNVs, Indels and Fusions

The HP2 assay performance on clinical cfDNA was assessed using a set of 137 plasma samples from 11 centers. The cfDNA mutational profile, representing a total of 388 genomic alterations (SNVs (N=322), Indels (N=50) and Fusions (N=16)) were validated with orthogonal NGS methods on FFPE or cfDNA samples (Supplementary Table S3). Samples were processed with the HP2 assay using 30 ng input DNA and sequenced on NextSeq 500/550/550Dx (Mid-output flow cell) or NovaSeq 6000 (SP flow cell) instruments (Supplementary Table S22). The SNVs, Indels and Fusions detection concordance with orthogonal methods was assessed against NGS and PCR/ddPCR (Table 3, Supplementary Table S23).

**Table 3:**
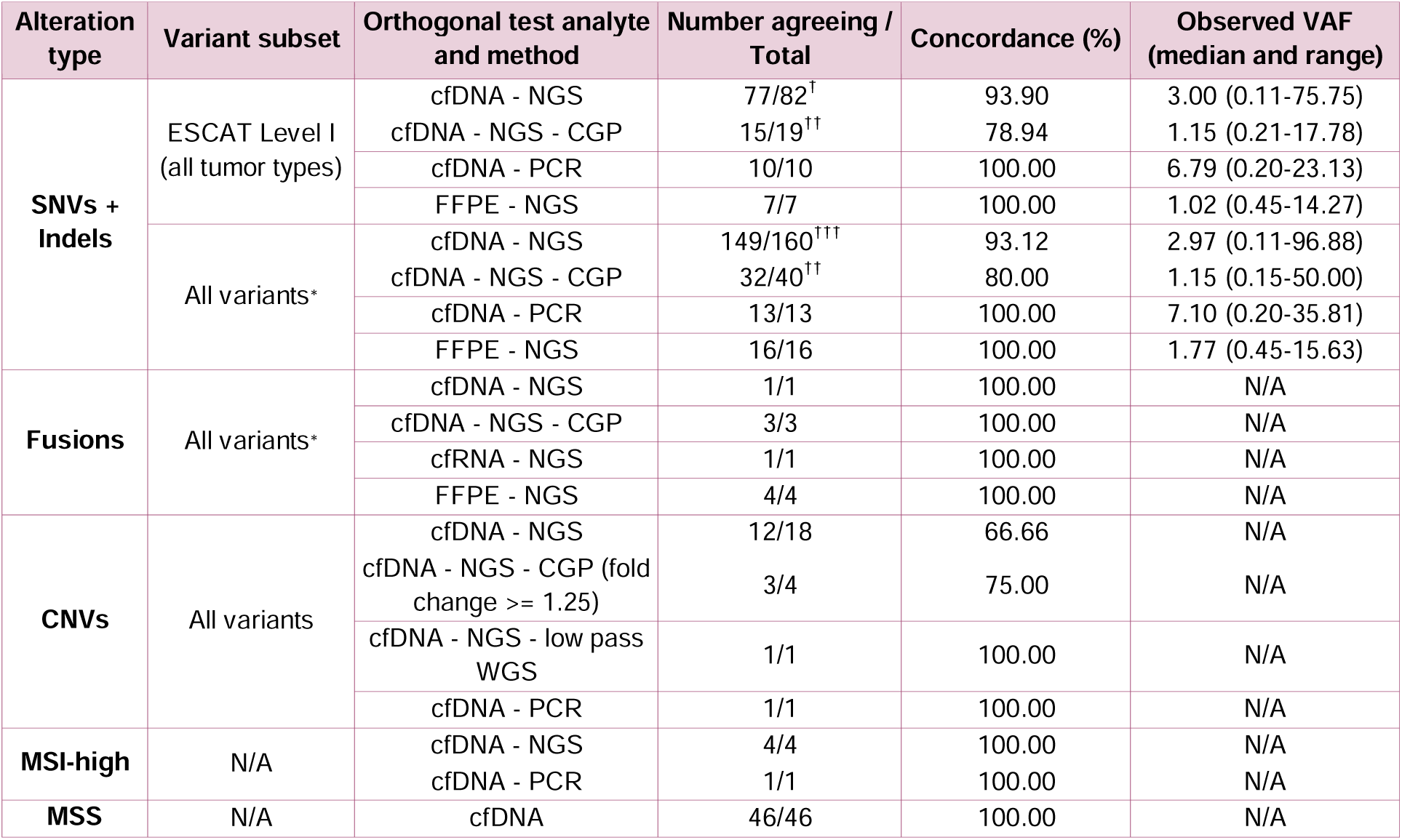
Evaluation of HP2 assay concordance with orthogonal methods in clinical cfDNA samples. SNVs, Indels, Fusions and CNVs detection, as well as MSI status determination concordance between HP2 and orthogonal methods, including NGS small panels on same cfDNA samples (cfDNA - NGS) or matching FFPE samples (FFPE - NGS), NGS large panels on same cfDNA samples (cfDNA - NGS - CGP), and PCR/ddPCR assays on same cfDNA samples (cfDNA - PCR). Data obtained on 125 clinical cfDNA samples. *: All variants with high clinical relevance, as defined by the following criteria: being classified as Pathogenic/Likely pathogenic in the ClinVar database, unlikely to be polymorphisms (<1.00% allele frequency in the gnomAD database) and being linked to relevant therapies. †: 1 disagreeing variant from a library preparation not passing QC criteria, ††: 3 disagreeing variants from library preparations with low cfDNA input (<10 ng), †††: 4 disagreeing variants from library preparations either not passing QC criteria or with low cfDNA input (<30 ng). (CGP= Comprehensive genomic profiling, WGS=Whole genome sequencing)

The impact of input DNA quantity on the HP2 sensitivity was assessed using a single clinical cfDNA sample with 30 ng, 20 ng and 10 ng of input DNA. The 2 orthogonally validated mutations included (at 12.70% and 2.10% VAF) were detected in all 3 tested conditions (Supplementary Table S24). The inter-instruments concordance was assessed using a set of 5 clinical cfDNA libraries (representing 6 SNVs/Indels and 1 MSI-high status) sequenced on both an Illumina NextSeq 500 (Mid output flow cell) and NovaSeq 6000 (SP flow cell) instruments. The observed VAF of SNVs and Indels were significantly correlated between instruments (R^2^=0.99) and the MSI status was consistently determined across runs (Table 4). It is expected to observe small variations in VAF between distinct assays, as differences in probe design and bioinformatics algorithms typically affect target capture efficiency and VAF estimation.

**Table 4:** Analytical performance of HP2 assay across instruments in clinical samples. Observed VAF% in clinical samples, sequenced using 30 ng of input DNA for HP2, across two different Illumina platforms. Comparative VAF% and input ng from other vendor kits are also detailed.

| Sample ID | Variant | Input DNA other vendor | Expected VAF other vendor | Observed VAF HP2 @30 ng input |  |
| --- | --- | --- | --- | --- | --- |
|  |  |  |  | NextSeq 500 | NovaSeq 6000 |
| Sample 1 | EGFR L858R | 36.8 ng | 0.23% | 0.38% | 0.68% |
| Sample 2 | EGFR E746_S752delinsA | 66.2 ng | 0.11% | 0.39% | 0.11% |
|  | EGFR<br>E746_S752delinsV |  | 85.51% | 40.91% | 40.99% |
| Sample 3 | KRAS Q61H | 36.8 ng | 1.50% | 3.00% | 3.00% |
| Sample 4 | KRAS G12C | 62.5 ng | 1.41% | 0.14% | 0.13% |
| Sample 29 | KRAS G12D | 26.0 ng | 3.50% | 3.64% | 3.49% |
|  | MSI-high |  | MSI -High | MSI -High<br>Score - 0.92<br>Affected Sites - 10 | MSI -High<br>Score - 0.99<br>Affected Sites - 11 |

In NGS assays, recovering sufficient amounts of cfDNA to ensure an optimal sensitivity is a typical challenge. The performance of two NGS assays requiring different input DNA quantities was compared using a set of 11 clinical cfDNA samples representing 16 SNVs and Indels, and one Fusion. DNA libraries were processed either with the HP2 assay, using 13.3 ng to 40 ng input DNA, or another commercially available NGS assay requiring 100 ng input DNA (37.7 ng to 100 ng DNA used). The HP2 assay detected more clinically actionable variants, while the alternative NGS LBx assay failed to detect 3 alterations despite using more ng input (Supplementary Table S25).

### CNV detection performance

The HP2 assay CNV detection performance was assessed on a set of cfDNA reference standards with CNVs at 0.5% and 2.0% spike-in frequencies (Fig. 2a, Table 1). Additional testing on FFPE reference standards (Seraseq® Compromised FFPE Tumor DNA Reference Material) showed a 100% sensitivity for both *MET* and *ERBB2* CNVs (4 total copies).

**Fig. 2:**
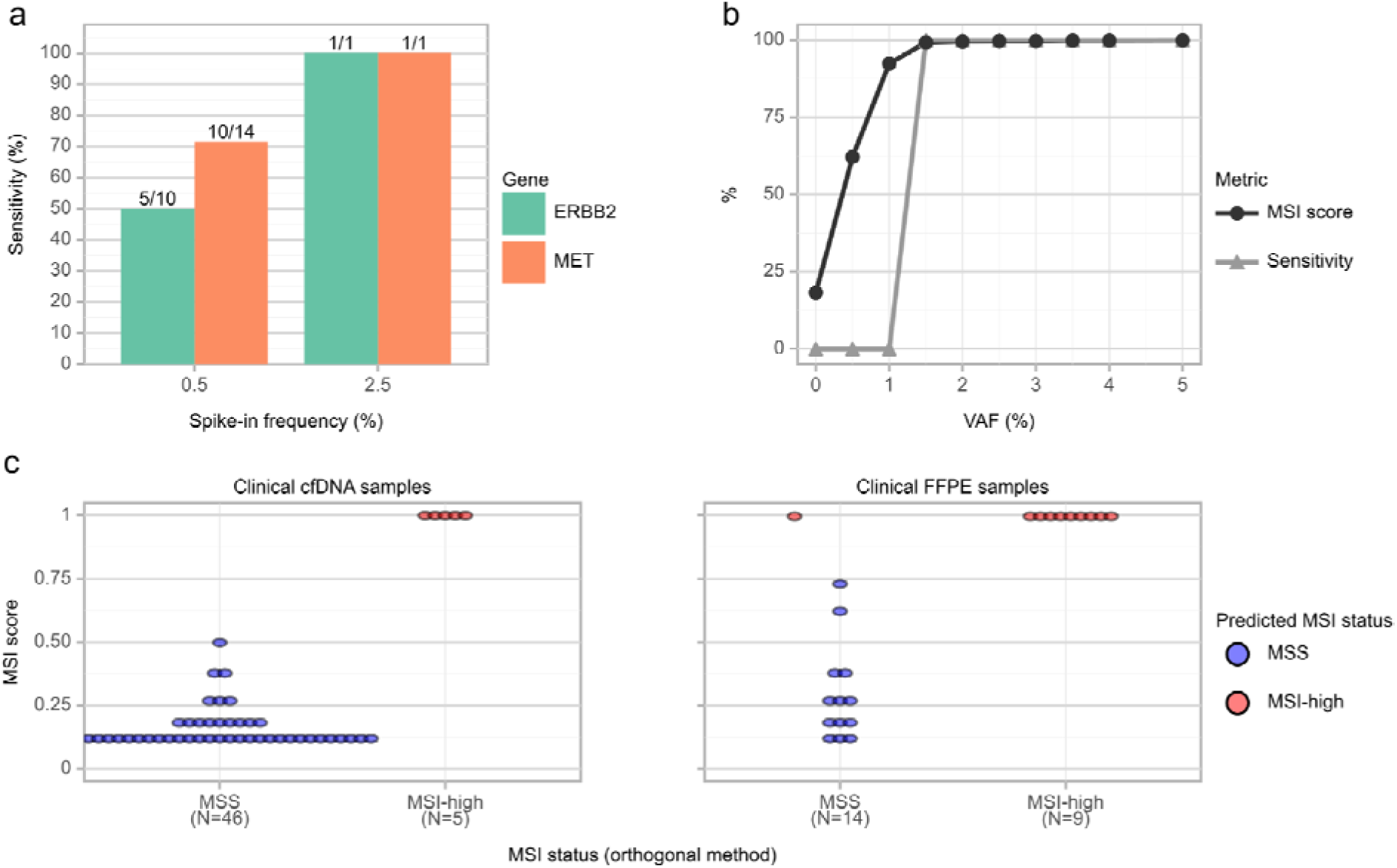
Analytical performance for CNV detection and MSI status determination. **a.** Analytical sensitivity for the detection of *MET* and *ERBB2* CNVs in reference standards at 0.5% and 2.5% spike-in frequency. **b.** MSI score reported by Hedera Prime and MSI status determination sensitivity on *in-silico* dilutions of reference standards. **c.** Estimated MSI score and predicted MSI status by HP2 versus predicted MSI status by orthogonal assays on clinical cfDNA samples and clinical FFPE samples.

Out of 137 available clinical cfDNA samples, 18 presented copy number gains reported by orthogonal assays, within genomic regions covered by HP2. Seventeen out of 24 copy number gains (70.83%) were concordantly identified by HP2 and orthogonal methods (Table 3). Tumors can be heterogeneous, and cfDNA may not fully represent all subclones present in the tumor, leading to discrepancies between different detection methods. And moreover each method has its own technical limitations and biases, which can contribute to differences in detection rates.

### MSI analytical performance

The HP2 assay performance for the determination of MSI status was evaluated on reference standards over a range of *in-silico* dilutions. The MSI status determination sensitivity was 100.00% in all tested VAFs above 1.0% and dropped to 0.00% below 1.0% VAF (Fig. 2b, Table 1). The estimated VAF of individual MSI sites varied largely. In order to conserve the diversity of the allele frequencies in the mixed samples and resembling an *in-vitro* dilution, a global mixture coefficient was applied on the entire sample and no correction was applied to account for site-specific variations.

The performance of the HP2 assay for determining the MSI status was assessed on 74 clinical samples from cancer patients (42 cases of NSCLC, 13 cases of CRC, 6 cases of Endometrial cancer, 7 samples of other cancer types) (Supplementary Table S26). Samples included 51 cfDNA and 23 FFPE (including 2 sequencing replicates) with orthogonally validated MSI status (9 MSI-high in total) along with four matched normal gDNA samples. The assay showed 100.00% and 95.83% concordance with orthogonal methods on cfDNA and FFPE respectively (Fig. 2c, Table 3).

A single FFPE clinical sample showed discrepancy in MSI classification between the HP2 panel and the orthogonal fragment analysis. HP2 identified 14 of 36 unstable sites, while already 6 unstable sites may lead to a MSI-high classification, resulting in a high MSI score of 0.993. In contrast, the orthogonal test with only five markers including BAT-25, NR-21, and NR-24 for which HP2 found strong evidence, reported stability. This highlights potential limitations in marker selection or sensitivity differences between these methods, with fragment analysis possibly missing instability signals detected by HP2.

## Discussion

Liquid biopsy (LBx) heralds a revolutionary advance in oncology, offering a repeatable molecular monitoring of patients to determine the genetic profile evolution of tumor via circulating materials such as cfDNA. This non-invasive and minimally innovative follow-up greatly facilitates dynamic monitoring of tumor genetic evolution, precise identification of somatic alterations, and real-time assessment of therapeutic response, all while minimizing patient morbidity (15,16). LBx enhances traditional tissue biopsies in several significant ways: it is a less burdensome procedure for patients, facilitates frequent sampling and is representative of the tumoral heterogeneity and inherently reduces the sampling bias associated with tissue biopsies (17). LBx‘s integration into precision medicine paradigms is transformative, affording timely and nuanced genetic insights that refine treatment strategies (18) increasing the likelihood of detecting actionable mutations and ultimately improving patient outcomes (19,20). Nonetheless, the field faces hurdles in augmenting assay accuracy and achieving inter-laboratory standardization before it can supplant large-panel NGS tissue genotyping. Additionally, despite validation studies demonstrating high sensitivity and specificity for current liquid biopsy assays, there is a paucity of corroborative studies using orthogonal methods or cross-platform NGS comparisons (21–23).

Herein, we highlight comprehensive validation study of the HP2 assay, which affirmed its robust technical performance, exhibiting high sensitivity and specificity for detecting SNVs, Indels, CNVs, Fusions, and MSI in reference materials. These outcomes demonstrated remarkable reproducibility across different runs and platforms. To further confirm clinical sensitivity of the HP2 assay, we assessed its performance on a cohort of 137 samples encompassing various cancer types sourced from multiple hospitals across different countries.

In reference standards, using 30 ng of cfDNA input material, the sensitivity for detecting SNVs and Indels at 0.5% VAF was 97.9% and 93%, respectively, with a combined specificity of 99.67% for ESCAT Level I (all tumor types). Technical performance for detecting Indels was slightly lower reflecting the inherent challenges in than SNVs and also complicating technical challenges in sequencing and bioinformatics pipelines which can lead to alignment and calling errors (24) especially in short-read sequencing data (150bp) and cfDNA reference standards which have an average DNA fragment length of 160bp, mirroring cfDNA (25) where the signal-to-noise ratio diminishes. Additionally, bioinformatics tools may not be optimized for low-frequency detection, leading to increased false positives or missed calls. These complexities underscore the need for continuous refinement of detection methodologies to improve accuracy in identifying Indels. Nevertheless, this level of performance is comparable to other high-sensitivity ctDNA assays reported in the literature, which typically achieve >90% sensitivity for variants at 0.5% VAF with high specificity (26).

For SNV/Indels, the study demonstrates a significant correlation between input DNA and variant detection sensitivity (Fig. 1e, Supplementary Table S20). At higher input (30ng), detection rates for SNVs and Indels were notably lower at 0.1% VAF, improving significantly at 0.25% VAF, particularly for Indels (Fig. 1d, Supplementary Table S18). In contrast, with lower input (10ng) but higher VAF (0.5%), detection rates were substantially higher for both SNVs (91%) and Indels (89%). These findings underscore the critical balance between input DNA and VAF thresholds in achieving optimal variant detection, highlighting the need for careful consideration of these parameters in assay design and interpretation.

In a recent study by Wenjin Li et al (2024) and Deveson IW et al (2021), nine commercially available ctDNA assays along with five benchmark assays were compared. These studies have reported sensitivities ranging from 33% to 80% for various Fusions. In regard to this, HP2 performance surpasses all other commercial methods as it achieved 100% sensitivity and specificity at 0.5% VAF using 30ng input DNA, with high sensitivity maintained even at lower VAFs (88.89% at 0.1% and 92.59% at 0.25%) (Fig. 1d, Supplementary Table S18). The assay’s robustness is evident in its ability to maintain high accuracy with reduced input DNA also (100% at 20ng and 92.59% at 10ng), addressing key challenges in LBx assays like detecting Fusions accurately with minimal input is crucial for clinical samples where DNA quantity is often limited. This capability is particularly valuable for monitoring minimal residual disease, early detection of recurrence, and tracking treatment response in cancer patients, especially in cases where Fusion genes play a crucial role in pathogenesis and treatment selection (27).

The HP2 assay’s performance in the QuIP proficiency test underscores its efficacy in detecting clinically significant *ESR1* and METex14 mutations, outperforming several commercial NGS LBx-specific assays. RNA-based methods have been favored for their ability to directly detect the skipped exon in transcripts. However, the success of HP2, a DNA-based method, in the QuIP proficiency test suggests that it has overcome the limitations typically associated with DNA-based detection of these complex mutations. This robust analytical validity suggests the HP2’s potential to enhance precision oncology in ER-positive breast cancer management particularly in guiding endocrine therapy decisions and detection of driver mutations in NSCLC. Further validation in diverse clinical contexts need to be implemented to solidify HP2 utility in routine diagnostic settings.

The study demonstrated high clinical concordance (94% for ESCAT Level I variants) compared to previous research (1,28), attributed to expansive gene coverage, CNV/MSI detection, and inclusion of more Fusions. Using the orthogonal cfDNA NGS method, we identified 18 discordant variants due to either VAFs below 0.1%, inadequate library QC, or insufficient input material (<30ng). NGS detection of ctDNA is constrained by the low frequency of mutant alleles in circulation and the limited amount of cfDNA available. Although NGS can quantify mutant alleles at ultra-deep frequencies, it necessitates substantial template cfDNA, intricate molecular barcoding, and extensive bioinformatics, which impede rapid clinical application (29). Additionally, the quality and quantity of cfDNA are compromised by its short half-life and potential genomic DNA contamination during blood collection (30). Detecting low-level variants is challenging due to the required high sequencing depth and the risk of false positives at very low VAFs (31). These discordant variants highlight the limitations of NGS when VAFs are low, library quality is suboptimal, or input material is insufficient.

Orthogonal analysis via ddPCR on a subset of samples (N=15) encompassing 35 variants provided additional validation, though three variants with VAFs below 0.1% were excluded to avoid bias. The superior sensitivity of ddPCR for low VAFs down to 0.1%, differs from NGS methodologies by utilizing higher input material and measured for single gene analysis (32). Additionally, three variants showed discordant results compared to tissue-based methods, likely due to temporal discrepancies in sample collection (Supplementary table S3). This discordance highlights the challenges in comparing plasma and tumor-tissue biopsies, influenced by factors such as tumor heterogeneity, morphology, and stage. The absence of a mutation in ctDNA present in tissue may reflect biological realities rather than poor analytical performance of the liquid biopsy assay (33). These findings underscore the complementary nature of tissue and liquid biopsies in comprehensive molecular profiling strategies for cancer patients, emphasizing the importance of considering both approaches for a more complete genomic landscape (19,20,34). This necessitates careful consideration when comparing results between these orthogonal assays.

The HP2 assay demonstrates exceptional accuracy in detecting MSI across diverse cancer types and sample sources, with 100% sensitivity for both cfDNA and FFPE samples. This aligns with recent advancements in cfDNA-based MSI detection, which have shown promising results in overcoming challenges associated with low tumor fraction and germline DNA contamination (35). The assay’s robust performance at VAFs above 1% highlights its potential for clinical applications. The high concordance between cfDNA and tissue MSI status in this study supports cfDNA MSI testing as a potential non-invasive alternative, consistent with large-scale studies reporting accurate identification of tissue MSI-H (87%) and MSS (99.5%) cases (36). While such results are promising, it’s important to note that factors such as tumor stage and vascular invasion may influence the sensitivity of cfDNA-based MSI detection (37).

The HP2 assay exhibits robust accuracy in CNV detection, addressing a critical challenge in clinical genomics. However, its lower sensitivity (62.5%) at 0.5% VAF in reference standards indicates limitations in detecting very low-frequency CNVs, a common issue in ctDNA-based methods (22,38). Factors affecting CNV calling in ctDNA include ctDNA concentration, experimental biases, and tumor heterogeneity (39,40). The assay’s initial results regarding sensitivity are very promising, with 2/2 events confirmed by gold-standard techniques like ddPCR and low-pass WGS accurately detected (41). The HP2 assay’s ability to detect CNVs without paired normal samples and its high specificity make it a valuable tool for CNV detection, potentially improving the efficiency and reliability of genomic profiling in oncology and beyond, especially in cases where tissue biopsies are impractical (42). The study’s scope precluded the inclusion of copy loss samples, constraining the assessment of detection sensitivities for these events. Accurately identifying copy loss remains complex because of low signal-to-noise ratios and the subtle nature of these alterations, a limitation noted in many CNV detection methodologies (43).

This study represents the inaugural analytical validation of an oncology LBx kit executed within a decentralized clinical framework; however, this approach is not without its limitations. The development of clinically valid ctDNA sequencing assays requires a multifaceted approach that bridges analytical and clinical validation. While contrived reference samples are invaluable for initial analytical performance assessment, they cannot fully recapitulate the complex biological factors encountered in real patient samples, such as the challenge of distinguishing tumor-derived mutations from benign variants arising from clonal hematopoiesis (22,44). Consequently, clinical thresholds like LOD and LOB must be established using patient-derived samples that encompass the full spectrum of biological variability. A stepwise validation process is crucial, starting with analytical validation using contrived samples, followed by preliminary testing with diverse clinical samples, refinement of assay parameters, and large-scale clinical validation studies (45). Ultimately, only through rigorous clinical trials can the true potential of ctDNA assays be realized, ensuring their effective translation from bench to bedside in the complex landscape of cancer diagnostics and monitoring (46).

## Conclusion

A recent ESMO study on the real-world availability of molecular technologies (47) revealed that, even in high-income countries across Europe, relevant NGS panels are still largely unavailable in routine clinical practice and are primarily confined to research settings. This limitation restricts patient access to targeted therapies and contributes to the significant gap between advances in anticancer drug development and the delivery of these treatments to patients (48).

The HP2 test panel represents a significant advancement that could help bridge this gap by enabling more patients to access targeted drugs. It facilitates tumor mutational profiling in liquid biopsies within decentralized laboratory settings, which are typical outside the US. HP2 can detect SNVs, Indels, Fusions, and CNVs with high sensitivity and specificity in 32 genes relevant to solid tumors and allows for MSI status characterization.

Its DNA-only workflow requires relatively small DNA input quantities and is compatible with Illumina sequencers, along with a dedicated data analysis platform, making it an efficient solution for hospital laboratories, pending approval for diagnostic use.

Unlike other large-scale NGS validations reported in the literature, this study is notable for its rigorous comparison against multiple independent NGS assays, conducted across a diverse array of clinical samples from 11 different institutions worldwide. To the author’s knowledge, this is the first reported analytical validation of an oncology LBx panel conducted in a decentralized clinical setting, incorporating an efficient and advantageous strategy alongside participation in proficiency testing programs and inter-laboratory comparisons, and provides valuable insights into the performance of the HP2 NGS LBx assay.

## Data Availability

All data produced in the present work are contained in the manuscript

## Acknowledgements

The authors would like to thank all patients and their families, the investigators, nurses, pathologists and the Hedera Dx team.

The authors are very grateful to AstraZeneca for their long-term collaboration with HCL Lyon-Sud Hospital. The authors thank AstraZeneca for their continued financial support of this analytical study.

Their gratitude also extends to the below two labs for participation in the inter-laboratory comparison study and who graciously agreed to publish their single sample data, contributing significantly to the robustness of the findings.

1. Diagnóstica Longwood, Av. Diagonal plaza., 40, 50197 Zaragoza, Spain
2. Mauri Keinänen, Department of Genetics, Fimlab Laboratories, Tampere, Finland.

## Authors’ contributions

LP, SM-B, TV, AR, and JB wrote the manuscript. LP, PC, GC, AH, MH, NP, E-MM, M-LK, LM-R, AD-L, GT, and ES provided edits to the first draft in manuscript writing. LP, PC, GC, EH, LM-R, AD-L, SM-B, TV, AR, JB, GR, FP, ES, HD, RvM, and ID analyzed the data. LP, PC, GC, EH, LM-R, AD-L, SM-B, TV, AR, JB, FP, UM, ES, HD, RvM, AD, and ID interpreted the data. GL, MP, AH, MH, HS, NP, E-MM, M-LK, A-HA, VB, RG, SM-B, RvM, HD, ID, and ES performed the HP2 and orthogonal assays. LP, MP, AH, MH, HS, PS, A-HA, VB, EH, LM-R, AD-L, ES, and AD contributed to the recruitment of patients.

All authors read and approved the final manuscript.

## Data availability

The datasets generated during and/or analyzed during the current study are available from the corresponding author on reasonable request.

## Ethics declarations

Under respective participating country regulations, there is an exception to the clinical research rules for method validation. In practice, the analysis of routine samples, i.e. samples not collected specifically for research purposes and without any change in patient management (or diagnosis), is outside the scope of research and therefore does not require an approach to the authorities or an ethics committee. Analysis of samples using a new kit against the gold standard does not require personal data (no personal data associated with the samples is processed in this context) and the samples are considered anonymous. This study is therefore outside the scope of the RGPD requirements as no personal data is processed. The study was conducted in full compliance with the principles outlined in the Declaration of Helsinki, ensuring ethical standards were maintained throughout the process.

Samples from University Clinical Hospital of Santiago de Compostela (Spain) are part of a collection associated with the protocol approved by the Ethics Committee of Santiago de Compostela and Lugo Ethics Committee (Ref: 2017/538). Written informed consent was obtained from every participant.

