## Supplementary material for "Analytical Validation of a Pan-Cancer NGS Assay for In-House Liquid Biopsy Testing: An International Multicenter Study": Legends of suplementary data

### Supplementary materials and methods

**cfDNA isolation from plasma:** cfDNA was isolated from a minimum of 4 mL of plasma using the MagMAXTM Cell-Free Total Nucleic Acid Isolation Kit (Applied Biosystems, Thermo Fisher Scientific, Waltham, MA, USA) or AVENIO cfDNA Isolation Kit V2 (Roche Diagnostics GmbH, Mannheim, Germany) or QIAamp Circulating Nucleic Acid Kit (Qiagen, Hilden, Germany) or the EZ1&2 ccfDNA Kit with EZ1 Advanced XL instrument (Qiagen, Hilden, Germany) or Maxwell RSC ccfDNA LV Plasma kit (Promega, Madison, WI, USA) according to the manufacturer's guidelines.

#### Bioinformatics pipeline and data analysis

**Genome Alignment:** The software performs read alignment against the hg19 reference genome using the BWA mem (Burrows-Wheeler Aligner) software.

**Duplex consensus and SNVs / Indels calling:** Based on UMI sequences and base quality score, individual reads are collapsed into single-strand and double-strands consensus reads (representing a single DNA fragment). Subsequently, the VarDict variant caller calls somatic variants on the duplex consensus reads. Following this, normalization of variants is conducted, and variants supported by only one read are discarded.

**CNV module:** The CNV module calculates the average coverage in a predefined set of target genomic regions and normalizes it within each sample to enable cross sample comparisons. A set of representative background samples are chosen to establish expected coverage levels per region. Amplified regions are identified via statistical testing, contrasting target region coverage against that observed in int the same regions within other samples. Employing a segmentation algorithm, contiguous regions are clustered into potential CNV events.

**Fusion module:** The de-novo DNA-based Fusion detection module extracts candidate breakpoint pairs from soft clipped reads in a predefined set of clinically relevant target regions. A binary classification model computes a score for each candidate breakpoint pair based on features extracted from UMIs, alignment quality, base quality, sequence complexity and length of supporting softclips as well as number of supporting reads. Fusions are then called based on a predefined score threshold. At most one breakpoint pair is reported for each pair of target regions. For the fusion module development and further training of the model, 30 cfDNA commercial reference samples were used independently, not overlapping with samples used for accuracy calculations.

**MSI module:** The Hedera Prime MSI module predicts the sample level MSI status based on aggregated results from 36 individual genomic simple repeat loci in which homopolymer length distribution profiles are calculated. These distributions are compared to background distributions of homopolymer profiles estimated on microsatellite stable samples. Significant deviations of these homopolymer length profiles are reported as candidate MSI signals. Samples may be classified as Microsatellite stable (MSS) or Microsatellite unstable (MSI-high) if at least 6 sites show statistically significant alterations from the expected repeat length. Taking into account the gradual nature of the MSI signal, the MSI module also reports an MSI intermediate state if at least 4 sites show statistically significant alterations. These thresholds are derived from comprehensive literature analysis, ensuring alignment with established proportions of MSI sites. This methodology not only adheres to scientific precedent but also enhances the reliability of MSI status determination (Gilson P et al., Cancers (Basel). 2021).

In addition to the classification result an MSI score is provided comprising results of all repeat loci in a single indicative value. For the MSI module development and further training of the model, 12 FFPE commercial reference samples were used independently, not overlapping with samples used for accuracy calculations.

**Data analysis:** Analytical sensitivity (a.k.a. recall or true positive rate), specificity (a.k.a. true negative rate), precision (a.k.a. positive predictive value, or PPV), concordance (a.k.a. percent agreement) and Limit of Detection (LoD) were calculated using the following formulas:

$Sensitivity = 100 x TP / (TP {+ FN)}$

$Specificity = 100 x TN / (TN {+ FP)}$

$Precision = 100 x TP / (TP {+ FP)}$

$Concordance = 100 x OA / (OA {+OD)}$

With:

*TP*: number of true positive variants

*FP*: number of false positive variants

*TN*: number of true negative variants (estimated as the number of bases included in the assay’s target minus the sum of *tp*, *fn* and *fp*)

*FN*: number of false negative variants

*OA*: number of observed agreements

*OD*: number of observed disagreements

LoD: The limit of detection is estimated as the spike-in frequency at which variants are detected with > 90% sensitivity.

All boxplots represent first and third quartiles, with the median as a middle line and whiskers at the last value within a 1.5×IQR distance respectively from the upper or lower quartile, where IQR is the interquartile range. Lines in all violin plots represent the first, second, and third quartiles.

#### Analytical validation

All analytical validation tests were conducted in a College of American Pathologists (CAP)-accredited and National Accreditation Board for Testing and Calibration Laboratories (NABL)-accredited laboratory.

**SNVs and Indels detection performance:** To assess the HP2 SNVs and Indels detection performance, we used a set of 36 DNA libraries derived from 4 cfDNA reference standards at 0.5% spike-in VAF (Twist cfDNA reference, SensID 5-Gene-Multiplex reference, Horizon Multiplex I reference and Horizon Structural Multiplex reference), including a total of 1348 SNVs and 802 Indels, and one negative control cfDNA standard (SensID 0% AF Ashkenazim Son. Each library was prepared using the HP2 kit with 30 ng input DNA and sequenced on an Illumina NextSeq 2000 instrument (P2 flow cell). The SNVs and Indels detection sensitivity was estimated using the 0.5% spike-in reference standards, while the corresponding specificity was estimated using the negative controls.

**Fusions detection performance:** To assess the HP2 Fusion detection performance, we used a set of 56 DNA libraries derived from 6 cfDNA reference standards, including a total of 54 fusions. These included 3 reference standards for fusions at 0.5% spike-in VAF (Horizon Structural Multiplex reference, Seraseq® ctDNA Complete™ Mutation Mix and Seraseq® ctDNA Mutation Mix v2) and 3 negative controls cfDNA standards (SensID 5-Gene-Multiplex reference, Horizon Multiplex I reference and SensID 0% AF Ashkenazim Son). Reference standards used to train the Hedera Prime fusion classification algorithm were not included in this validation set. Each library was prepared using the HP2 kit with 30 ng input DNA, and sequenced either on an Illumina NextSeq or NovaSeq 6000 instrument. The gene fusion detection sensitivity and specificity were estimated using all the included libraries.

**CNV detection performance:** To assess performance of HP2 on CNVs, we used a set of 15 DNA libraries derived from 2 cfDNA reference standards, including a total of 26 CNVs. These included 2 reference standards for CNVs at 0.5% and 2% spike-in VAF (Horizon Structural Multiplex reference and Seraseq® ctDNA Complete™ Mutation Mix). Each library was prepared using the HP2 kit with 30 ng input DNA, and sequenced either on an Illumina NextSeq or NovaSeq 6000 instrument. The CNV detection sensitivity and specificity were estimated using all the included libraries.

**MSI status determination performance:** To assess the performance of HP2 for the determination of the MSI status, we used a set of 2 DNA libraries derived from the SensID MSI tumour sample with 16% and 0% allele frequencies. The sensitivity of the MSI prediction was evaluated on a range of *in-silico* dilutions mimicking MSI signals at allele frequencies from 0% to 5% (binomial sampling on reads including the same total number of reads in each dilution).

**Inter- and intra-assay reproducibility:** The reproducibility of the HP2 assay was assessed on a set of 30 DNA libraries derived from 4 cfDNA reference standards at 0.5% spike-in VAF (Twist cfDNA reference, SensID 5-Gene-Multiplex reference, Horizon Multiplex I reference and Horizon Structural Multiplex reference), including a total of 862 SNVs and Indels, and one negative control cfDNA standard (SensID 0% AF Ashkenazim Son). Two identical sequencing libraries were prepared independently (by different persons on different days) from the same set of reference standards using the HP2 kit with 30 ng input DNA. The two libraries were sequenced in independent runs on an Illumina NextSeq 2000 instrument with P2 flow cell. The SNVs and Indels detection sensitivity was estimated using the 0.5% spike-in reference standards, while the corresponding specificity was estimated using the negative controls. Concordance between runs was estimated as the percentage of variants consistently detected across the two sequencing runs, assuming individual DNA libraries are all independent. Concordance within runs was estimated as the percentage of variants consistently detected across two DNA libraries derived from the same reference standard.

**Inter-laboratory reproducibility:** The reproducibility of the HP2 assay between independent diagnostic laboratories was assessed using one cfDNA reference standard including 8 SNVs and Indels (SensID 5-Gene-Multiplex reference). The libraries were diluted at 5.0% (undiluted) and 1.0% (5x dilution) spike-in frequency. Identical sequencing libraries were prepared and sequenced independently in each laboratory using 30 ng input DNA. Libraries were sequenced either on an Illumina NextSeq 550 (Mid-output flow cell) or NextSeq 2000 (P2 flow cell) instrument.

**Inter-instrument reproducibility:** To assess the reproducibility of the HP2 assay with different sequencing instrument models, we used a set of 15 DNA libraries derived from 5 cfDNA reference standards. These included 4 reference standards for fusions at 0.5% spike-in VAF (Twist cfDNA reference, SensID 5-Gene-Multiplex reference, Horizon Multiplex I reference and Horizon Structural Multiplex reference), including a total of 862 SNVs and Indels, and one negative control cfDNA standard (SensID 0% AF Ashkenazim Son). Each library was prepared using the HP2 kit with 30 ng input DNA. The same libraries were sequenced on 3 runs, each with a different instrument (NextSeq 550 with Mid-output flow cell, NextSeq 2000 with P2 flow cell and NovaSeq 6000 with SP flow cell). The SNVs and Indels detection sensitivity was estimated using the 0.5% spike-in reference standards, while the corresponding specificity was estimated using the negative controls. Concordance was estimated as the percentage of variants consistently detected across sequencing runs on different instruments (i.e. a variant is concordant if it is detected in the same DNA library). The concordance was reported both between all pairs of runs and across all runs.

**Impact of VAF on variant detection:** To assess the impact of VAF on the detection performance of the HP2 assay, we used a set of DNA libraries derived from 2 cfDNA reference standards (Twist cfDNA reference and SensID 5-Gene-Multiplex reference), with spike-in frequencies at 0.1%, 0.25%, 0.5%, 1%, 2.5% and 5% (representing 36 DNA libraries in total) (Supplementary Table S1). Each library was prepared using the HP2 kit with 30 ng input DNA, and sequenced either on an Illumina NextSeq 2000 (P2 flow cell) or NovaSeq 6000 (SP flow cell) instrument.

**Impact of input DNA quantity on detection performance:** To assess the impact of the input DNA quantity on the detection performance of the HP2 assay, we used a set of DNA libraries derived from 1 cfDNA reference standard (Twist cfDNA reference), with spike-in frequencies at 0.5% and input DNA quantities of 10 ng, 20 ng, 25 ng, 30 ng and 50 ng (representing 15 DNA libraries in total) (Supplementary Table S1). Each library was prepared using the HP2 kit and sequenced either on an Illumina NextSeq or NovaSeq 6000 instrument.

#### Supplementary tables

Supplementary tables are provided in the separate ‘supplementary_tables.xlsx’ file:

- Table S1: List of commercially available reference standards.
- Table S2: Distribution of diagnosed tumor types in clinical samples.
- Table S3: List of clinical cfDNA samples and orthogonal variant detection methods.
- Table S4: List of clinical FFPE samples and orthogonal variant detection methods.
- Table S5: Hedera Profiling 2 ctDNA test panel target regions. Ex: Exon , CDS: Coding sequence, ECD: extracellular domain.
- Table S6: Overview of key sequencing metrics obtained on reference standards and clinical cfDNA samples used for the HP2 assay analytical validation.
- Table S7: Overview of key sequencing metrics obtained on reference standards and clinical cfDNA samples used for the comparison of coverage.
- Table S8: Mean coverage obtained in reference standards and clinical samples.
- Table S9: Overview of key sequencing metrics obtained on reference standards used to assess SNVs, Indels and Fusions detection performance.
- Table S10: Summary of SNVs, Indels and Fusions detection performance per individual variant.
- Table S11: Detailed SNVs and Indels detection performance data.
- Table S12: Overview of key sequencing metrics obtained on reference standards used to assess inter- and intra-assay concordance.
- Table S13: inter- and intra-assay concordance. Percentage of variants consistently detected between two sequencing runs (inter-assay concordance) and between replicates derived from the same reference standard (intra-assay concordance).
- Table S14: Overview of key sequencing metrics obtained on reference standards used to assess inter- and inter-instrument reproducibility.
- Table S15: Summary of SNVs, Indels and Fusions detection performance obtained across multiple sequencing instruments.
- Table S16: Inter-instrument concordance.
- Table S17: Overview of key sequencing metrics obtained on reference standards used to assess the impact of VAF on performance.
- Table S18: Impact of VAF on performance. Summary of HP2 analytical sensitivity on SNVs, Indels and fusions at varying spike-in frequencies.
- Table S19: Overview of key sequencing metrics obtained on reference standards used to assess the impact of input cfDNA quantity on performance.
- Table S20: Impact of input cfDNA quantity on performance. Summary of HP2 assay analytical sensitivity on SNVs, Indels and fusions at varying input DNA quantities.
- Table S21: QuIP 2024 - ESR1 Liquid biopsy proficiency test summary.
- Table S22: Overview of key sequencing metrics obtained on clinical cfDNA samples.
- Table S23: Summary of variant detection agreement between HP2 and orthogonal methods in clinical cfDNA. Data shown for variants classified as ESCAT Level 1.
- Table S24: Analytical performance as a function of varying cfDNA input for a clinical sample.
- Table S25: Comparison of HP2 analytical performance with another NGS ctDNA kit vendor for clinical samples using different ng input cfDNA.
- Table S26: MSI detection concordance with orthogonal methods on clinical cfDNA and FFPE samples.

#### Supplementary figures

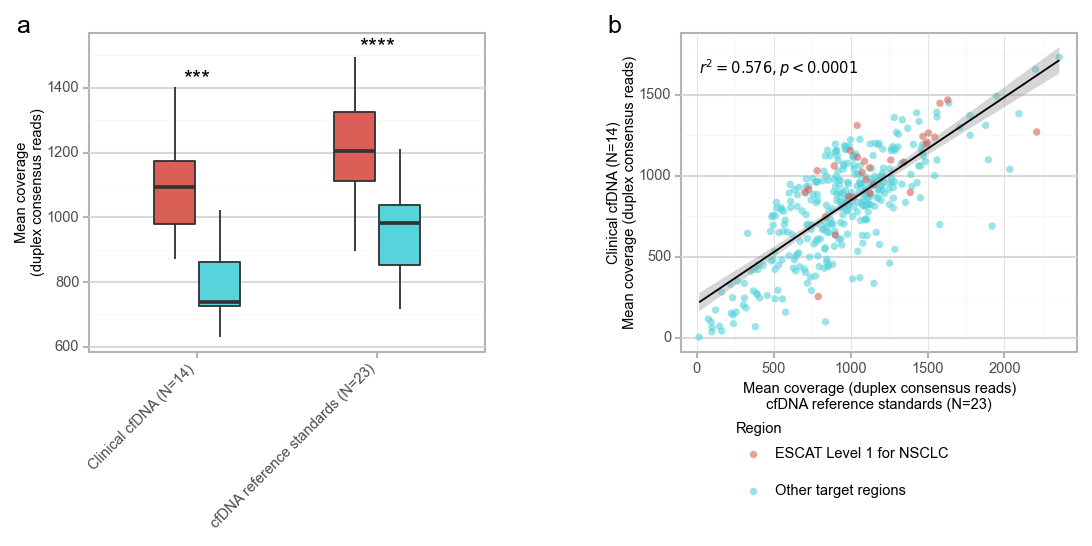

**Fig. S1: Observed coverage in reference standards and clinical cfDNA. a.** Mean duplex consensus reads coverage per gene exon covered by the HP2 panel. Data shown separately for exons relevant for ESCAT Level 1 mutations in NSCLC and the rest of the panel target. Statistical significance assessed by Wilcoxon test. ***: p-value<0.001, ***: p-value<0.0001. **b.** Comparison of mean coverage (per region bins of size ~150bp) obtained on clinical cfDNA and cfDNA reference standards. Correlation assessed with Python's scipy.stats.pearsonr method.

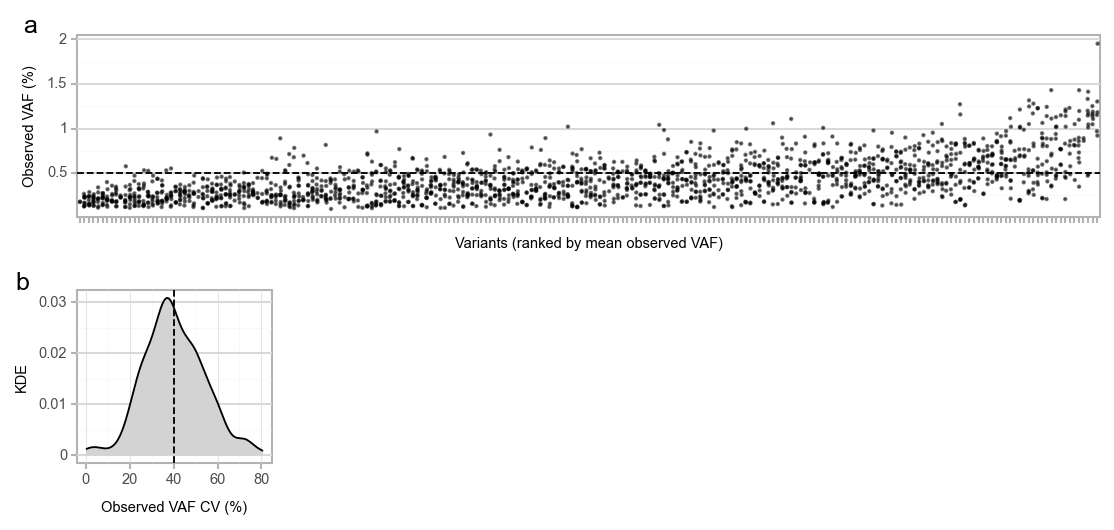

**Fig. S2: Observed VAF in reference standards. a** Observed VAF per detected variant in reference standards at 0.5% spike-in frequency. **b** Kernel density estimate (KDE) showing the distribution of observed VAF coefficient of variation in reference standards at 0.5% spike-in frequency.Dashed line represents the mean.

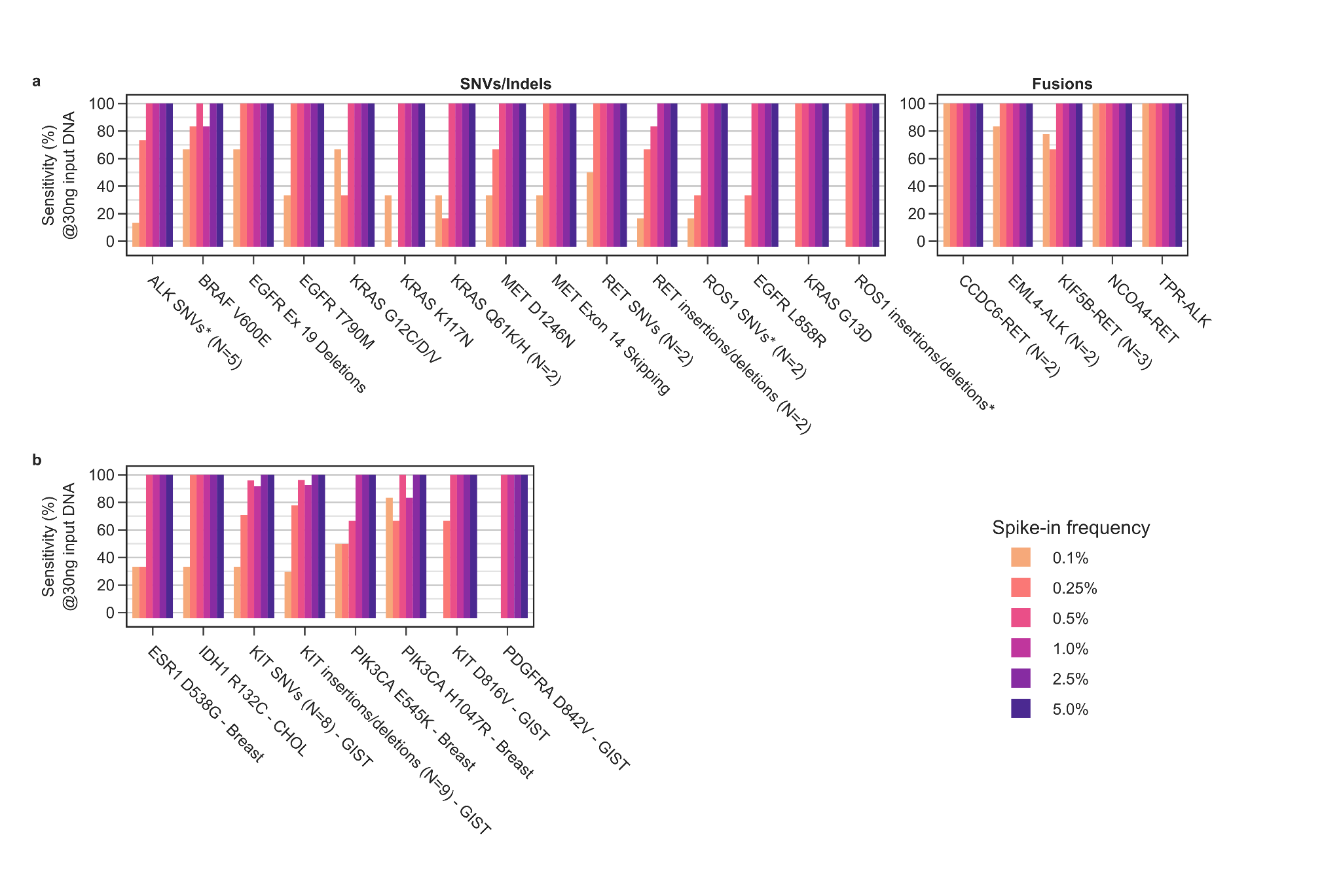

**Fig. S3: Impact of VAF for the detection of key genomic alterations.** HP2 assay analytical sensitivity for SNVs, Indels and fusions at varying spike-in frequencies. Data shown (**a**) for tested variants classified as ESCAT level of evidence 1 for NSCLC, and (**b**) for tested variants classified as ESCAT level of evidence 1 for any tumor type (except NSCLC only). *:Acquired resistance kinase domain mutations.

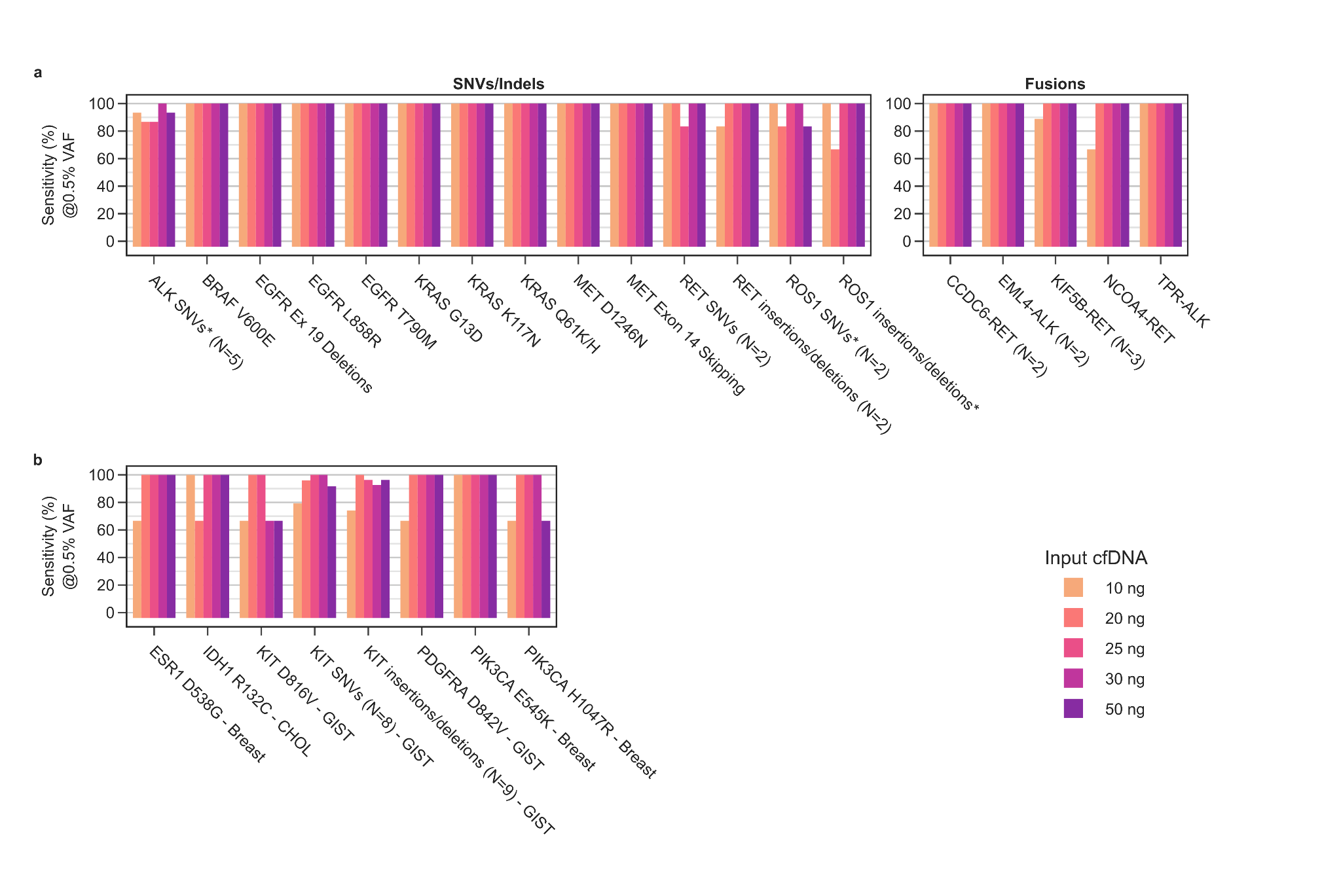

**Fig. S4: Impact of input DNA quantity for the detection of key genomic alterations.** HP2 assay analytical sensitivity on key genomic alterations at varying input DNA quantities. **a** Tested SNVs, Indels and fusions classified as ESCAT Level 1 genomic alterations for NSCLC. **b** Tested SNVs and Indels classified as ESCAT Level 1 genomic alterations for other tumor types. Results obtained on reference standards at 0.5% spike-in frequency. *: Acquired resistance kinase domain mutations.

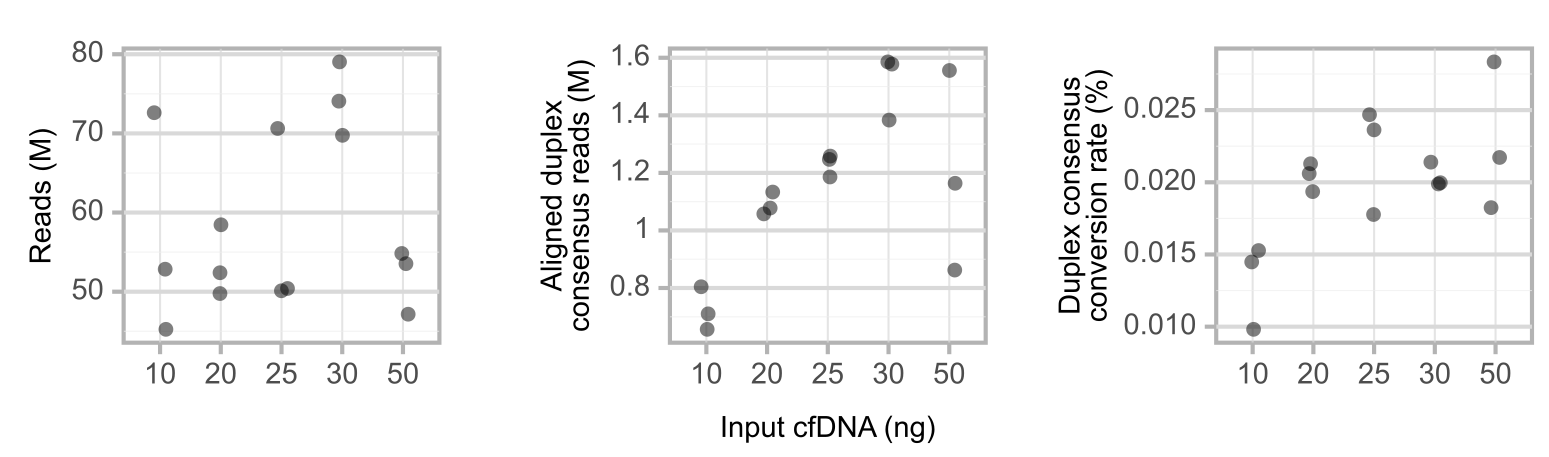

**Fig. S5: Impact of input DNA quantity on sequencing reads.** Number of sequencing reads and called duplex consensus reads obtained from DNA libraries prepared from different input DNA quantities. The duplex consensus conversion rate is calculated as the number of aligned duplex consensus reads divided by the number of reads.
